# A dataset for evaluating clinical research claims in large language models

**DOI:** 10.1101/2024.10.08.24315103

**Authors:** Boya Zhang, Anthony Yazdani, Alban Bornet, Philipp Khlebnikov, Marija Milutinovic, Hossein Rouhizadeh, Poorya Amini, Douglas Teodoro

## Abstract

Large language models (LLMs) have the potential to enhance the verification of health claims. However, issues with hallucination and comprehension of logical statements require these models to be closely scrutinized in healthcare applications. We introduce CliniFact, a scientific claim dataset created from hypothesis testing results in clinical research, covering 992 unique interventions for 22 disease categories. The dataset used study arms and interventions, primary outcome measures, and results from clinical trials to derive and label clinical research claims. These claims were then linked to supporting information describing clinical trial results in scientific publications. CliniFact contains 1,970 scientific claims from 992 unique clinical trials related to 1,540 unique publications. Intrinsic evaluation yields a Cohen’s Kappa score of 0.83, indicating strong inter-annotator agreement. In extrinsic evaluations, discriminative LLMs, such as PubMedBERT, achieved 81% accuracy and 79% F1-score, outperforming generative LLMs, such as Llama3-70B, which reached 52% accuracy and 39% F1-score. Our results demonstrate the potential of CliniFact as a benchmark for evaluating LLM performance in clinical research claim verification.

## 1. Background & Summary

Large language models (LLMs) have demonstrated remarkable success in several natural language processing tasks in the health and life sciences domain^1^. Due to parameter scaling, access to specialized corpora, and better human alignment techniques, performance has significantly improved in recent years^2^. Yet, they still struggle with factual accuracy in various domains^3^. LLMs may produce factual errors that contradict established knowledge available at the time^4^. These inaccuracies and errors are particularly concerning in critical fields like healthcare, where incorrect information can have severe consequences^5^.

To mitigate issues with factual accuracy and vulnerability to hallucinations, the incorporation of domain-specific knowledge when evaluating LLMs has been proposed^6^. This stems from the fact that factual accuracy^7^ and vulnerability to hallucinations^8^ in LLMs can vary significantly across domains^9^. Models fine-tuned for a general purpose tend to outperform in the general domain^6^ while models fine-tuned for specific domains, such as medicine (e.g., Meditron^10^, Med-PaLM^11^), often outperform general-purpose models in those areas.

Another critical challenge for LLMs is their ability to perform logical reasoning^12^. This is particularly important in clinical research, where scientific claims are posed as logical statements, such as ‘the intervention *X* is more effective than placebo for a specific outcome’^13^, that are either *true* or *false*. Evaluating these claims requires a strong understanding of hypothesis testing and causal inference^14^. However, the nature of LLMs, which are trained to predict tokens within a context^15^, makes them struggle with complex logical statements^16^, even making unfaithful reasoning^17^.

Research has shown that LLMs can be easily misled by irrelevant information^18^. Chain-of-thought (CoT) prompting can improve multi-step reasoning by providing intermediate rationales^19^. Concerns remain regarding the faithfulness and reliability of these explanations, as they can often be biased or misleading^20^. Furthermore, while methodologies such as self-correction can improve reasoning accuracy, current models still struggle to correct their errors autonomously without external feedback^21^. In some cases, their performance degrades after self-correction^21^. Integrating LLMs with symbolic solvers for logical reasoning^22^ and hypothesis testing prompting for improved deductive reasoning^23^ are proposed to address these limitations.

Claim verification datasets play a crucial role in assessing the factual accuracy of LLMs across various domains^24^. FEVER^25^, a general-domain dataset, was created by rewriting Wikipedia sentences into atomic claims, which are then verified using Wikipedia’s textual knowledge base. FEVER also introduces a three-step fact verification process: document retrieval, evidence selection, and stance detection. In the political domain, the UKP Snopes corpus^26^, derived from the Snopes fact-checking website, includes 6,422 validated claims paired with evidence text snippets. For the scientific domain, SciFact^27^ includes 1.4K expert-written biomedical scientific claims paired with evidence containing abstracts annotated with labels and rationales while Climate-FEVER^28^ contains 1,535 claims sourced from web searches, with corresponding evidence from Wikipedia.

Specifically to the health and life science domains, PUBHEALTH^29^ gathers public health claims from fact-checking websites and verifies them against news articles. ManConCorpus^30^ contains claims and sentences from 259 abstracts linked to 24 systematic reviews on cardiovascular disease. The COVID-19 pandemic and its infodemic effect^7,31^ have further motivated the development of specialized datasets. HealthVer^32^ is a medical-domain dataset derived by rewriting responses to questions from TREC-COVID^33^, verified against the CORD19 corpus^34^. Similarly, COVID-Fact^35^ targets COVID-19 claims by scraping content from Reddit and verifying them against scientific papers and documents retrieved via Google search. CoVERt^36^ enhances claim verification in the clinical domain by providing a new COVID verification dataset containing 15 PICO-encoded drug claims and 96 abstracts, each accompanied by one evidence sentence as rationale. These datasets are either focused on lay claims^29,32,35^, which require simpler reasoning skills, or, when focused on complex clinical research claims, they are disease-specific, e.g., COVID-19^36^ or cardiovascular^30^ and of reduced scale (*O*(10^1^) claims)^30,36^. Thus, they are limited to evaluating the factuality of complex clinical research claims by LLMs.

To reduce this gap, we propose CliniFact, a large-scale claim dataset to evaluate the generalizability of LLMs in comprehending factuality and logical statements in clinical research. CliniFact claims were automatically extracted from clinical trial protocols and results available from ClinicalTrials.gov. The claims were linked to supporting information in scientific publications available in Medline, with evidence provided at the abstract level. The resulting dataset contains *O*(10^3^) claims spanning across 20 disease classes. This new benchmark offers a novel approach to evaluating LLMs in the health and life science domains, with specific challenges to understanding claims at the logical reasoning and hypothesis testing levels.

## 2. Methods

We utilized the ClinicalTrials.gov database as our primary data source, which comprises an extensive collection of registered clinical trials and their respective results. From each selected clinical trial, we systematically extracted key components to create the research claim, including the primary outcome measure, intervention, comparator, and type of statistical test. These components form the basis for generating one or more claims from each trial. We used the corresponding PubMed abstracts linked to these clinical trials as evidence to make judgments on the claims. In the following, we detail the dataset construction process.

### 2.1 Resources

The dataset uses information from two resources maintained by the U.S. National Library of Medicine: ClinicalTrials.gov and PubMed. ClinicalTrials.gov is a comprehensive online database that provides up-to-date information on clinical research studies and their results. These clinical trials serve as the most reliable medical evidence for evaluating the efficacy of single or multiple clinical interventions^37^. PubMed primarily includes the MEDLINE database of references and abstracts on life sciences and biomedical topics. The empirical evidence for clinical trial outcomes is often described in the results of clinical research studies published in medical journals indexed by PubMed^38^.

### 2.3 Data acquisition and pre-processing

On January 15^th^, 2024, we downloaded a total number of 57,422 clinical trials from CT.gov (https://clinicaltrials.gov/search) that met the following criteria: *i*) Study Status: *Terminated* or *Completed*; ii) Study Type: *Interventional*; and *iii*) Study Results: *With results*. The resulting dataset is a compressed zip file containing individual raw JSON files of each study named by the clinical trial identifier (NCTID).

### 2.4 Dataset construction

In the following, we formally describe the claim generation process. An overview of the pipeline is illustrated in Figure 1, and an example of the extracted fields is illustrated in Table 1.

**Table 1:** Example of the extracted fields in CliniFact.

|  |  |
| --- | --- |
| **NCTID** | NCT00234065 |
| **PMID** | 20833591 |
| **Outcome Title** | Numbers of Patients With First Occurence of Stroke |
| **Outcome Time Frame** | From start of treatment to end of follow-up period ( follow-up periods : 29 months [Standard Deviation 16, range 1-59 months]) |
| **Intervention** | Cilostazol |
| **Comparator** | Aspirin |
| **Type of Statistical Test** | Non-Inferiority or Equivalence |
| **Publication Title** | Cilostazol for prevention of secondary stroke (CSPS 2): an aspirin-controlled, double-blind, randomised non-inferiority trial. |
| **Publication Abstract** | [...] Interpretation: Cilostazol seems to be non-inferior, and might be superior, to aspirin for prevention of stroke after an ischaemic stroke, and was associated with fewer haemorrhagic events. Therefore, cilostazol could be used for prevention of stroke in patients with non-cardioembolic stroke. [...] |
| **Hypothesis Label** | Evidence |

**Figure 1:**
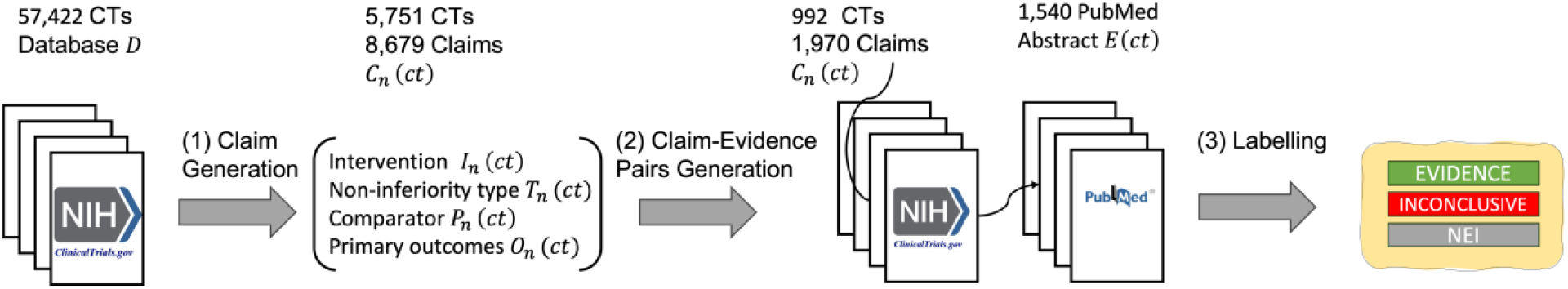
Overview of the CliniFact dataset construction pipeline with three major modules: claim extraction, claim-evidence pairing, and labeling.

#### Claim extraction

Let *D* represent the filtered ClinicalTrials.gov database we downloaded, with each clinical trial represented as *ct*′ *ϵ D*. From the *ct*′ set, we extracted the intervention, outcome measures, and comparator information from the subset of clinical trials *ct* limiting the selection to trials with bi-arm groups (see details in Section 2.4.1) and reporting p-value results for the primary outcome measures. For the ct set, we then extracted the primary outcome measures *O*_*n*_ (*ct*), with their corresponding intervention *I*_*n*_ (*ct*), comparator *P*_*n*_ (*ct*), and type of statistical test *T*_*n*_ (*ct*). These components are utilized to construct one or more claims *C*_*n*_ (*ct*) for each clinical trial, where n can vary between 0 and N. We represent the generation of the claim *C*_*n*_ (*ct*) as function *f*, such that:

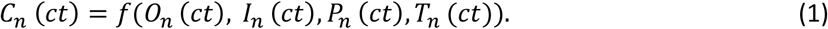

#### Claim-evidence pairing

For each ct *ϵ D*, there may be an associated scientific abstract *E*(*ct*) from PubMed reported by the authors of the study in ClinicalTrials.gov. We link *E*(*ct*) to each corresponding claim *C*_*n*_ (*ct*) to create a claim-evidence pair (*C*_*n*_ (*ct*), *E*(*ct*)). In this context, *E*(*ct*) represents the evidence from the PubMed abstract used to make judgements on the claim *C*_*n*_ (*ct*). An *E*(*ct*) might have two statuses depending on the type of information it provides: i) *background*, if it provides background information to the clinical trial, and ii) *result*, if it describes results for the clinical trial. This information is provided by the study authors and will be used further to label the claim-evidence pairs (*C*_*n*_ (*ct*), *E*(*ct*)).

#### Claim-evidence labeling

The labeling process for the claim-evidence pairs involves two key steps. First, for each claim *C*_*n*_ (*ct*), we assign a positive or negative label *L*_1_(*C*_*n*_ (*ct*)) based on the p-value reported for its respective primary outcome measure. A positive label is assigned if the p-value indicates a statistically significant result, while a negative label is assigned if the p-value indicates a lack of statistical significance. Following conventional statistical thresholds, we considered p-value < 0.05 as statistically significant. Second, we consider the link nature between the clinical trial and the scientific abstract. If a clinical trial *ct* is linked to a scientific abstract *E*(*ct*) that reports *results* for the trial, we further filter these instances to include only those where exactly one abstract is linked to the clinical trial. For these cases, the label for the claim-evidence pair *L*_2_(*C*_*n*_ (*ct*),*E*(*ct*)) is defined as “evidence” if the claim *C*_*n*_ (*ct*) is positive, and “inconclusive” if the claim *C*_*n*_ (*ct*) is negative. Conversely, if the scientific abstract *E*(*ct*) linked to the clinical trial provides *background* information, we include all the linked abstracts, and the label for the claim-evidence pair *L*_2_(*C*_*n*_ (*ct*), *E*(*ct*)) is defined as “not enough information” (NEI).

#### 2.4.1 Primary outcome-arm group pairs for claim generation

To extract primary outcome measures and arm group information from the clinical trial database *D*, we focused exclusively on clinical trials *ct ϵ D* that included bi-arm groups of types *Experimental* and *Comparator*. In a clinical trial, an arm refers to a group of participants that receives a particular intervention, treatment, or no intervention according to the trial’s protocol^39^. The arm type represents the role of each arm in the clinical trial. For generating the clinical research claim, we used the term *intervention* to represent the *Experimental* arm group and *comparator* to represent the *Comparator* arm group, with mappings provided in Supplementary Table S1. The intervention and comparator terminologies are grounded in the PICO framework, i.e., population, intervention, comparator, and outcome^40^.

In the study design section, arm groups are labeled as *Experimental* and *Comparator*, but these labels are not in the result section. The titles of arm groups also vary between the two sections. Thus, to label the arm groups in the result section as *intervention* or *comparator*, we followed the approach proposed by Shi *et al*.^41^. In this approach, we mapped arm group titles in the result section to the most similar one in the study design section by calculating the cosine similarity of their embeddings created using BioBERT^42^.

##### Efficacy label

We extract the efficacy label *L*_1_(*C*_*n*_ (*ct*)) for the primary outcome *O*_*n*_ (*ct*) from the measure analysis in the outcome measure information module. Each primary outcome *O*_*n*_ (*ct*) paired with arm groups *I*_*n*_ (*ct*), *P*_*n*_ (*ct*) may have one or multiple associated analyses, some of which include p-values representing the statistical significance of the results. We only extracted the analyses with p-values and compiled the p-values into a list for each primary outcome-arm group pair. We assigned a *positive* efficacy label to an outcome-group pair if any p-value in its associated list is smaller than 0.05, indicating statistical significance. We assigned a *negative* efficacy label if all p-values were equal to or greater than 0.05, indicating statistical non-significance. We extracted 8,679 primary outcome-arm group pairs, of which 4,179 were labeled as positive and 4,500 as negative, for 5,751 unique clinical trials.

#### Type of statistical test

Each primary outcome-arm group pair could be associated with a type of statistical test. We categorize these types into the ones outlined in the study data structure of ClinicalTrials.gov^43^. The outlined types include *Superiority, Noninferiority, Equivalence, Noninferiority or Equivalence*, and *Superiority or Other*. A *Superiority* test evaluates if a new treatment is better than another (e.g., standard treatment or placebo) by rejecting the null hypothesis of no difference^44^. A *Noninferiority* test shows that the new treatment is not significantly worse than the existing one, within a predefined margin^44^. An *Equivalence* test demonstrates that two treatments are statistically equivalent, with differences falling within a clinically insignificant margin^44^. *Noninferiority or Equivalence* tests first establish noninferiority, and then assess equivalence^45^. Lastly, *Superiority or Other* tests may also evaluate noninferiority or equivalence if superiority is not shown^46^.

#### Clinical research claim

A scientific claim is a verifiable statement. The claim should be atomic (about one aspect of the statement), decontextualized (understandable without additional context), and check-worthy (the veracity can be confirmed)^47^. In natural language, a clinical research claim can be expressed as a scientific claim in a format of ‘<Intervention> is <Type of Statistical Test> to <Comparator> in terms of <Outcome>.’ For example, for study NCT00234065 shown in Table 1, we could reframe the outcome-group pair to the following claim *C*_*n*_ (*ct*): Cilostazol is Non-Inferior or Equivalent to Aspirin in terms of Numbers of Patients With First Occurrence of Stroke From start of treatment to end of follow-up period (follow-up periods: 29 months [Standard Deviation 16, range 1-59 months]). The *Intervention* and *Comparator* terms are sourced from the *Arm/Group Title*, while the *Outcome* is the combination of *Outcome Measure Title* and *Outcome Measure Time Frame* available in the clinical trial protocol (Table 2).

**Table 2:** Fields extracted for clinical research claim generation.

| Section | Definition | Path |
| --- | --- | --- |
| Study Identification | Clinical Trial Identifier | A.nctId |
| Arm Groups and Interventions | Arm Type | B.armGroups.type |
|  | Intervention Type | B.interventions.type |
| Outcome Measure Information | Arm/Group Title | C.groups.title |
|  | Arm Description | C.groups.description |
|  | Type of Statistical Test | C.analyses.nonInferiorityType |
|  | Outcome Measure Type | C.type |
|  | Outcome Measure Title | C.title |
|  | Outcome Measure Time<br>Frame | C.timeFrame |
|  | Estimated Value | C.analyses.paramValue |
|  | Method | C.analyses.statisticalMethod |

Study Identification: A = protocolSection.identificationModule
Arm Groups and Interventions: B = protocolSection.armsInterventionsModule
Outcome Measure Information: C = resultsSection.outcomeMeasuresModule.outcomeMeasures

#### 2.4.2 Clinical trial-publication linkage

Publications corresponding to clinical trials were identified by their PubMed Identifiers (PMIDs) provided in the *Publications* section of the clinical trial results and categorized by reference types. As shown in Table 3, we created a CSV file detailing the clinical trial-publication relationships by extracting the NTC ID, PMID, and *reference type* from the filtered ClinicalTrials.gov database *ct* ∈ *D*. A total of 1,550 clinical trial-publication links were used in the balanced dataset, including 868 *background* links and 682 *results* links.

**Table 3:**
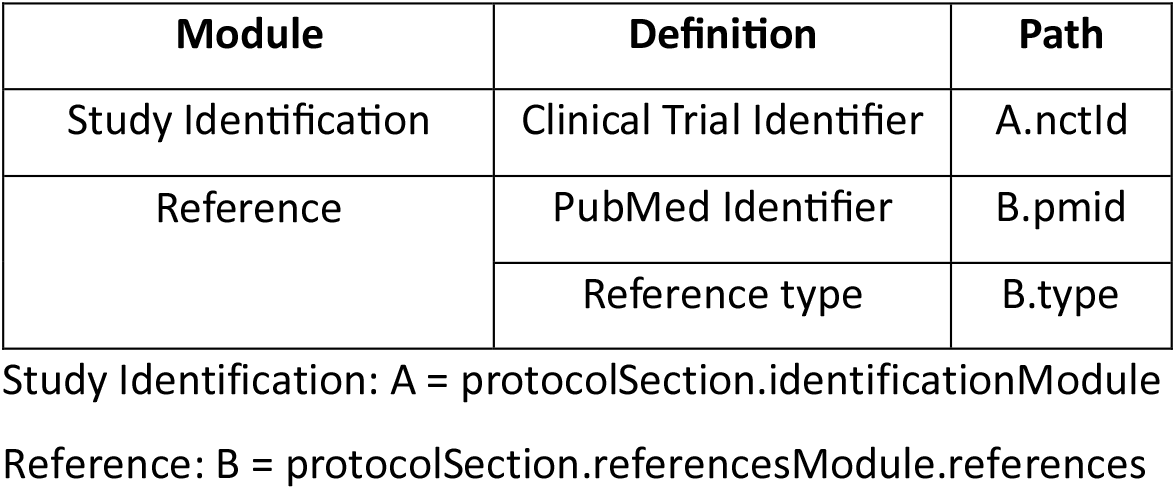
Fields used to create clinical trial-publication pairs.

| Module | Definition | Path |
| --- | --- | --- |
| Study Identification | Clinical Trial Identifier | A.nctId |
| Reference | PubMed Identifier | B.pmid |
|  | Reference type | B.type |

Study Identification: A = protocolSection.identificationModule
Reference: B = protocolSection.referencesModule.references

#### 2.4.3. Claim-evidence pairs generation

Using the extracted relationships between clinical trials and scientific publications, we linked claims to their corresponding publications to generate claim-evidence pairs. Each clinical trial may correspond to one or multiple publications categorized as either *background* or results. For *results* publications, we focused on clinical trials linked to a single publication. In these cases, if a claim *C*_*n*_ (*ct*) had a *positive* label, we labeled the claim-evidence pair as *evidence*. Conversely, if the claim had a *negative* label, we labeled the claim-evidence pair as *inconclusive*. For *background* publications, we labeled the claim-evidence pair as *NEI* regardless of whether the clinical trial was linked to one or multiple *background* publications. Using the PubMed API, we downloaded titles and abstracts of publications using PubMed unique IDs (PMIDs). We excluded samples with incomplete abstracts that are less than 15 words. The statistics for the number of words for the extracted primary outcomes and publications are illustrated in Table 4.

**Table 4:** Number of words for the text fields.

| Text Fields | Mean | Min | Max |
| --- | --- | --- | --- |
| Outcome Title | 9 | 1 | 38 |
| Outcome Time Frame | 7 | 1 | 43 |
| Publication Title | 16 | 2 | 68 |
| Publication Abstract | 247 | 22 | 1,013 |

## 3. Data Records

The dataset included 570 evidence, 415 inconclusive, and 8,196 not enough information (NEI) claim-evidence pairs. We further balanced the class distribution by down sampling the *NEI* class to 985 samples to match the total number of *evidence* and *inconclusive* samples. Our final dataset contains 1,970 primary claim-evidence pairs of 992 unique clinical trials and 1,540 unique publications. We show examples of *evidence, inconclusive* and *NEI* paired clinical research claims and abstracts in Table 5. The distribution of labels in the train, validation, and test splits is provided in Table 6.

**Table 5:** Examples of *evidence, inconclusive* and *not enough information* (NE) paired clinical research claims and abstracts.

| Claim | Publication title + abstract | Label |
| --- | --- | --- |
| Cilostazol is Non-Inferior or Equivalent to Aspirin in terms of Numbers of Patients With First Occurrence of Stroke From start of treatment to end of follow-up period (follow-up periods: 29 months [Standard Deviation 16, range 1-59 months]). | [...] Interpretation: Cilostazol seems to be non-inferior, and might be superior, to aspirin for prevention of stroke after an ischaemic stroke, and was associated with fewer haemorrhagic events. Therefore, cilostazol could be used for prevention of stroke in patients with non-cardioembolic stroke. [...] | **Evidence** |
| Nerve Block is Superior or Other to Periarticular Injection in terms of Post-Operative Pain Afternoon on post-operative Day 1, approximately 14:00. | [...] RESULTS: Mean pain scores on the afternoon of POD 1 were not different between groups (PNB group: 2.9 [SD 2.4]; PAI group: 3.0 [SD 2.2]; 95% confidence interval, -0.8 to 0.6; p=0.76). Mean | **Inconclusive** |
|  | pain scores taken at three times points on POD 1 were also similar between groups. [...] |  |
| Topiramate is Superior or Other to Placebo in terms of Percent Drinking Days (%DD) Weekly, weeks 1-12, average. | Combined pharmacotherapies and behavioral interventions for alcohol dependence: the COMBINE study: a randomized controlled trial. [...] | **NEI** |

**Table 6:** Distribution of labels in train, validation, and test splits.

| Set | Evidence | Inconclusive | NEI | All |
| --- | --- | --- | --- | --- |
| Train | 366 | 262 | 632 | 1,260 |
| Validation | 83 | 67 | 166 | 316 |
| Test | 121 | 86 | 187 | 394 |
| All | 570 | 415 | 985 | 1,970 |
| % | 29 | 21 | 50 | 100 |

In Figure 2, we show the clinical research claims stratified by the studied condition. Using the MeSH annotations provided by ClinicalTrials.gov, clinical research claims were associated with disease classes using the MeSH tree code (3 digits). In total, 20 disease categories (out of the 27 categories available in MeSH) are included in our dataset. It is important to note that a single clinical trial may be mapped to multiple disease classes according to the MeSH terminology. Therefore, when reporting the number of clinical research claims per disease, the total sample count may exceed the number of claims.

**Figure 2:**
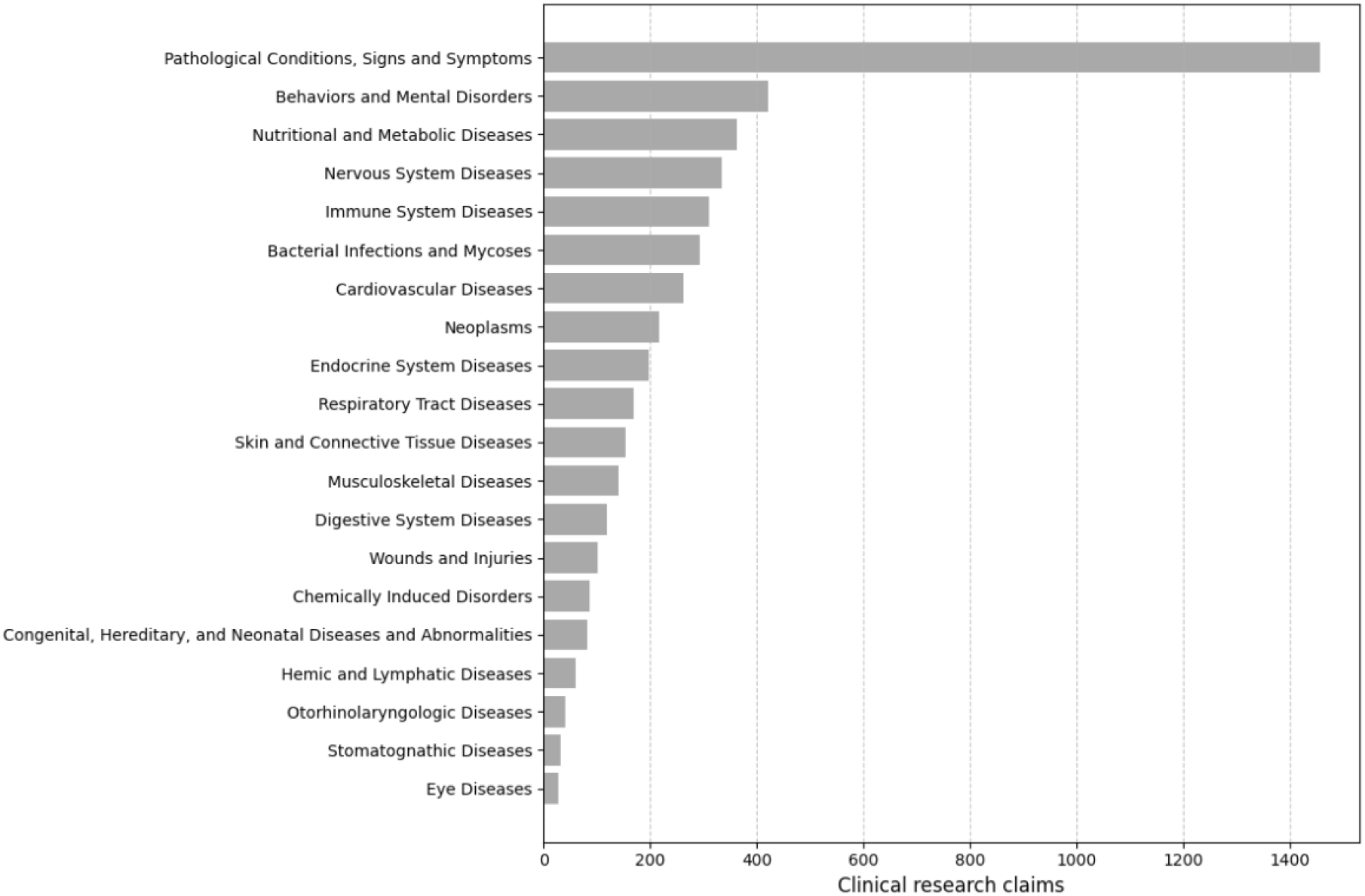
Clinical research claims stratified by study condition.

## 4. Technical Validation

### 4.1 Extrinsic evaluation

Given the claim *C*_*n*_ (*ct*) and the abstract *E*(*ct*), we investigated the performance of several discriminative and generative LLM to predict the label *L*_2_(*C*_*n*_ (*ct*), *E*(*ct*)) for the claim-evidence pair. We treated it as a multiclass classification problem, where the output indicates whether the abstract states that there is *evidence* for the claim, that it is *inconclusive*, or that the abstract does not provide information for the claim (*NEI*). For the discriminative LLMs, we concatenated a claim *C*_*n*_ (*ct*) and its corresponding abstract *E*(*ct*) with the special token [SEP] to form an input sequence [CLS, *C*_*n*_ (*ct*), SEP, *E*(*ct*)] and fed this input to the LLM. The model takes a sequence of tokens with a maximum length of 512 and produces a 768-dimensional sequence representation vector. For input shorter than 512 tokens, we added paddings (empty tokens) to the end of the text to make up the length. For input longer than 512 tokens, we truncated the abstract *E*(*ct*) from the beginning to make the input sequence fit into the 512 tokens. We provided the truncation algorithm in Supplementary Figure S1. For the generative LLMs, we concatenated a claim *C*_*n*_(*ct*) and its corresponding abstract *E*(*ct*) with the prompt shown in Table 7. In the zero-shot approach, we computed the probability of generating the token TRUE, FALSE, or NONE, and the token with the highest probability was the response. We fine-tuned the generative LLMs on the training split, evaluated their performance on the validation split, and selected the model with the lowest cross-entropy loss.

**Table 7:** Prompt for generative language models.

|  |  |
| --- | --- |
| **Instruction** | Given a scientific claim and an abstract, determine if the abstract reports positive results (TRUE), inconclusive results (FALSE), or offers no information (NONE) about the claim.<br>The task is to classify the pair claim abstract as follows:<br>TRUE: if the abstract provides positive support for the claim.<br>FALSE: if the abstract provides negative or inconclusive support for the claim.<br>NONE: if the abstract provides contextual or background information without directly reporting results about the claim. |
| **Input** | Claim: [...]<br>Abstract: [...] |
| **Response** | [...] |

We show the results of discriminative and generative LLMs on the test split in Table 8. The fine-tuned discriminative LLMs outperformed zero-shot and fine-tuned generative LLMs in the clinical research claim assessment. Specifically, PubMedBERT achieved the highest accuracy of 81.0% and an F1-macro score of 78.8%, showing improved effectiveness in processing biomedical text (p-value < 0.001, McNemar-Bowker Test), likely due to its domain-specific training. Other discriminative models like BioBERT and RoBERTa also performed well, with 78.2% and 77.4% accuracy, respectively. In contrast, zero-shot generative LLMs exhibited significantly lower performance, with OpenBioLLM-8B achieving the highest at 42.1% accuracy and an F1-macro of 30.5%, indicating limited capability in assessing biomedical claims without task-specific fine-tuning. Upon fine-tuning, generative LLMs showed notable improvements; for instance, Llama3-70B’s accuracy increased from 35.5% to 51.8%, and its F1-macro score from 23.0% to 39.7% (p-value < 0.001, McNemar-Bowker Test). Similarly, OpenBioLLM-70B improved from 35.0% to 49.7% accuracy after fine-tuning (p-value < 0.001, McNemar-Bowker Test). Nevertheless, they remain sub-optimal as compared to discriminative LLMs, despite a much higher number of parameters.

**Table 8:** Results of discriminative and generative language models on the test split.

| Training approach | Model | Accuracy (%) | F1-macro (%) |
| --- | --- | --- | --- |
| **Discriminative (fine-tuned)** | BERT Base (Uncased) | 72.6 | 69.0 |
|  | RoBERTa Base | 77.4 | 73.3 |
|  | BioBERT Base v1.1 (Cased) | 78.2 | 73.8 |
|  | PubMedBERT Base (Uncased) | **81.0** | **78.8** |
|  | MPNet Base v1 (QA) | 74.9 | 70.9 |
| **Generative (zero-shot)** | Llama3-8B | 33.2 | 19.8 |
|  | Llama3-70B | 35.5 | 23.0 |
|  | Meditron-7B | 30.7 | 15.7 |
|  | Meditron-70B | 33.2 | 22.9 |
|  | OpenBioLLM-8B | <u>42.1</u> | <u>30.5</u> |
|  | OpenBioLLM-70B | 35.0 | 28.3 |
| **Generative (fine-tuned)** | Llama3-8B | 38.3 | 26.9 |
|  | Llama3-70B | <u>51.8</u> | 38.7 |
|  | Meditron-7B | 30.7 | 15.7 |
|  | Meditron-70B | 30.7 | 15.7 |
|  | OpenBioLLM-8B | 45.4 | 34.5 |
|  | OpenBioLLM-70B | 49.7 | <u>39.7</u> |

We illustrate a detailed comparison of precision, recall, and F1-macro between the best discriminative and generative LLMs - PubMedBERT and OpenBioLLM-70 - across the classes *evidence, inconclusive*, and *NEI* in Figure 3(a). PubMedBERT consistently demonstrated superior and balanced performance across all metrics and classes. In contrast, OpenBioLLM-70B exhibited significant variability in its performance. The class *evidence* has a high recall of 86.8% but a lower precision of 39.5%, resulting in an F1-score of 54.3%. This result shows that the OpenBioLLM-70B, although finetuned, tends to predict *evidence* labels.

**Figure 3:**
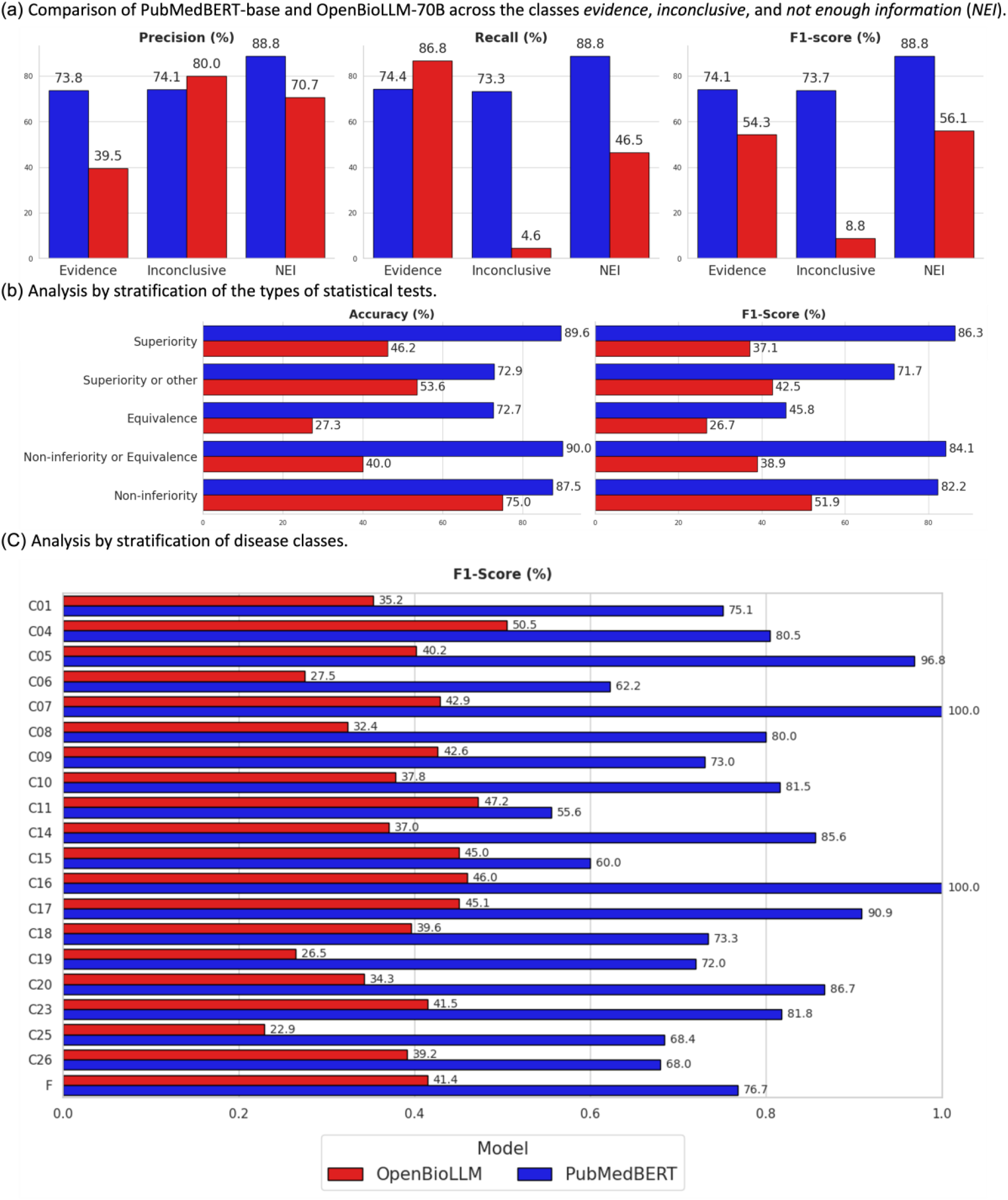
Performance and stratified analysis of the best discriminative and generative models.

Figure 3(b) shows the analysis of the best discriminative and generative LLMs across samples with different statistical test types. PubMedBERT consistently outperformed OpenBioLLM-70B in all categories and achieved its highest scores in the Non-inferiority or Equivalence (Accuracy: 90.0%, F1-macro: 84.1%) and Superiority (Accuracy: 89.6%, F1-macro: 86.3%) categories. Equivalence tests were the most challenging, with a performance of only 45.8% F1-macro. In contrast, OpenBioLLM-70B showed lower performance across all the types of statistical test, with its best results in the Non-inferiority category (Accuracy: 75.0%, F1-macro: 51.9%).

Figure 3(c) illustrates the performance of the leading discriminative and generative LLMs across 22 disease categories. PubMedBERT consistently outperformed OpenBioLLM in most disease classes. PubMedBERT’s best performance was in the disease category “Congenital, Hereditary, and Neonatal Diseases and Abnormalities” (C16), where it achieved a perfect F1-macro of 100%. Conversely, its lowest performance is in “Eye Diseases” (C11), with an F1-macro of 55.6%. OpenBioLLM-70’s highest F1-macro of 50.5% was achieved in the “Neoplasms” (C04) category. Its weakest performance occurred in the “Chemically Induced Disorders” (C25) class, where it recorded an F1-macro of 22.9%.

### 4.2 Intrinsic evaluation

To assess the intrinsic performance of our models, we conducted an evaluation using the subset from the CliniFact test set. Two researchers manually annotated 53 instances. We observed strong inter-annotator agreement (Cohen’s Kappa score = 0.83), confirming the quality of the dataset annotations. The best performing model (PubMedBERT) exhibited high agreement with the human annotators (Cohen’s Kappa scores of 0.74 and 0.70). Moreover, the model achieved an accuracy of 85% and 83%, with corresponding F1-macro scores of 79% and 74%.

## Usage Notes

CliniFact provides a benchmark for evaluating the accuracy of large language models (LLMs) in verifying scientific claims specific to clinical research. Researchers can utilize the dataset to develop and fine-tune models aimed at improving natural language understanding, logical reasoning, and misinformation^48^ detection in healthcare. Additionally, the dataset facilitates the comparison of performance across various types of LLMs.

## Data Availability

All data produced are available online at

https://github.com/ds4dh/CliniFact

## Code Availability

The entire process, from developing the CliniFact dataset to conducting experiments, was carried out using the Python programming language. The complete code and dataset are available on https://github.com/ds4dh/CliniFact.

## Author contributions

B.Z., P.A. and D.T. conceptualized the study, and B.Z. implemented the codes for the creation and evaluation of the dataset. P.K. and B.Z. performed human annotation. The manuscript was drafted by B.Z. and edited by A.B., A.Y. and D.T. All authors reviewed the final version.

