## Supplementary tables and figures for "A dataset for evaluating clinical research claims in large language models"

**Table S1:** Mapping arm types to intervention or comparator.

| Mapped Term | Arm Type |
|---|---|
| Intervention | Experimental |
| Comparator | Active Comparator |
| | Placebo Comparator |
| | Sham Comparator |
| | No Intervention |
| | Other |

**Figure S1:** Flowchart of the truncation algorithm.

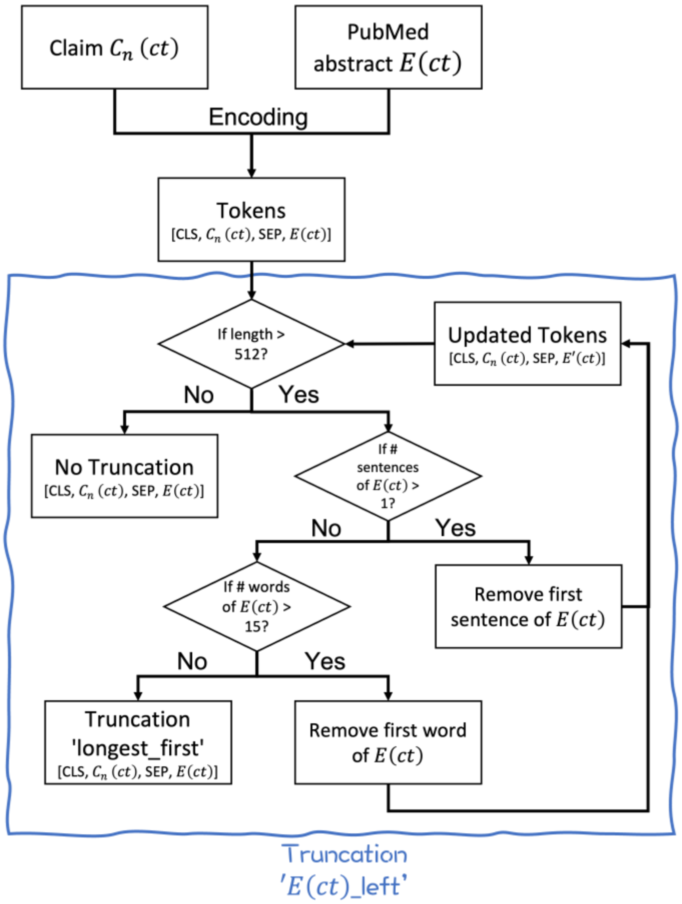
